# SARS-CoV-2 variants of concern dominate in Lahore, Pakistan in April 2021

**DOI:** 10.1101/2021.06.04.21258352

**Authors:** Muhammad Bilal Sarwar, Muhammad Yasir, Nabil-Fareed Alikhan, Nadeem Afzal, Leonardo de Oliveira Martins, Thanh Le Viet, Alexander J Trotter, Sophie J Prosolek, Gemma L Kay, Ebenezer Foster-Nyarko, Steven Rudder, David J Baker, Sidra-tul-muntaha, Muhammad Roman, Mark A Webber, Almina Shafiq, Balqees Shabir, Javed Akram, Andrew J Page, Shah Jahan

## Abstract

**Background:** The SARS-CoV-2 pandemic continues to expand globally, with case numbers rising in many areas of the world, including the Indian sub-continent. Pakistan has one of the world ‘s largest population, of over 200 million people and is experiencing a severe third wave of infections caused by SARS-CoV-2 that begun in March 2021.In Pakistan, during third wave until now only 12 SARS-CoV-2 genomes have been collected and among these 9 are from Islamabad. This highlights the need for more genome sequencing to allow surveillance of variants in circulation. In fact more genomes are available among travellers with a travel history from Pakistan, than from within the country itself.

**Methods:** For a better understanding of the circulating variants in Lahore and surrounding areas with a combined population of 11.1 million, within a week of April 2021, 102 samples were sequenced. The samples were randomly collected from 2 hospitals with a diagnostic polymerase chain reaction (PCR) cutoff value of less than 25 cycles.

**Results:** Analysis of the lineages shows that B.1.1.7 (first identified in the UK, Alpha variant) dominates, accounting for 97.9% (97/99) of cases, with B.1.351 (first identified in South Africa, Beta variant) accounting for 2.0% (2/99) of cases. No other lineages were observed.

**Discussion:** In depth analysis of the B.1.1.7 lineages indicates multiple separate introductions and subsequent establishment within the region. Eight samples were identical to genomes observed in Europe (7 UK, 1 Switzerland), indicating recent transmission. Genomes of other samples show evidence that these have evolved, indicating sustained transmission over a period of time either within Pakistan or other countries with low density genome sequencing. Vaccines remain effective against B.1.1.7, however the low level of B.1.351 against which some vaccines are less effective demonstrates the requirement for continued prospective genomic surveillance.

## Introduction

The COVID-19 pandemic has spread rapidly throughout the world and continues to expand in many regions. It began with an unknown case of pneumonia in the city of Wuhan, China (Huang *et al*., 2020). The causative pathogen has since been named ‘severe acute respiratory syndrome-related coronavirus 2 (SARS-CoV-2) ‘. As of May 2021, there have been over 167 million reported cases and 3.4 million fatalities (Dong, Du and Gardner, 2020). Genomic surveillance has assisted the pandemic response providing information for outbreak investigations and detecting possible epitope changes that would allow the virus to escape vaccines. Multiple classification systems have been developed to quickly communicate SARS-CoV-2 variants circulating in a community (Alm *et al*., 2020; Hodcroft *et al*., 2020; Rambaut *et al*., 2020). From these definitions, certain lineages have been designated as Variants of Concern (VOCs), which are defined as such due to indications of increased transmission patterns and/or possible resistance to vaccine and/or other treatments (Volz *et al*., 2021). World Health Organisation (WHO) has introduced a new nomenclature of these VOCs and Variants of Interest (VOIs) based on Greek alphabets.

B.1.1.7 (Alpha) and B.1.351 (Beta) are two VOCs that have circulated globally. The SARS-CoV-2 lineage B.1.1.7, designated Variant of Concern 202012/01 (VOC) by Public Health England, was first identified in the UK in late Summer to early Autumn 2020 (Volz *et al*., 2021). B.1.351 is another VOC identified in South Africa and defined by eight mutations in the spike protein including (K417N, E484K and N501Y) (Tegally *et al*., 2021).

Pakistan is currently experiencing a severe third wave of infections caused by SARS-CoV-2 which began in March 2021. Vaccination rates nationally are under 2% (*National Command Operation Center*, 2021), leaving large segments of the community at risk of serious illness from COVID-19. Currently very few SARS-CoV-2 genomes collected in Pakistan are available, with just 12 covering the third wave, 9 of which are from one city, Islamabad. This highlights the need for more genome sequencing from Pakistan, particularly given the current situation and the very high population in order to allow surveillance of variants in circulation. Currently, more genomes are available for travellers with a travel history from Pakistan, than from within the country itself (Shu and McCauley, 2017).

We have amplicon sequenced 102 samples, randomly chosen from a 1 week period in April 2021 from Lahore and surrounding areas, to get a snapshot assessment of the circulating lineage in the region. This has identified the B.1.1.7 variant of concern (Alpha) as the primary lineage circulating, found in 97.9% of cases, with clear signals of repeated overseas introductions into the region.

## Results and Discussion

In this study, from 2021-04-06 to 2021-04-11, 102 SARS-CoV-2 samples in viral transport medium (VTM) were randomly selected from SARS-CoV-2 diagnostic positives by the Mayo Hospital and Sheikh Zayed Hospital in Lahore, Pakistan. All samples had a Ct < 25. These public hospitals serve a predominantly middle income population. The patients were between 21 and 91 years of age with a broad distribution of ages, with 74% (n=74) male, 26% (n=26) female and 2 unknown. Four samples were from patients who died (all aged over 60), while all others recovered. Patients had no history of international travel, with 77 patients explicitly reporting that they and their household did not recently travel overseas (Supplementary Table S1).

Viral RNA was amplified using the ARTIC protocol (Quick, 2020) with sequencing libraries prepared using CoronaHiT (Baker *et al*., 2021). Resulting sequenced reads were used to generate consensus sequences with the ARTIC bioinformatics protocol (See methods).

Analysis of the PANGO lineages shows that B.1.1.7 (first identified in the UK) dominates, accounting for 97.9% (97/99) of cases, with B.1.351 (first identified in South Africa) accounting for 2.0% (2/99) of cases. No other lineages were observed.

SARS-CoV-2 was first identified in Pakistan on 26th February 2020 (Javed *et al*., 2020). The first B.1.1.7 genome was submitted to GISAID on 2020-12-25. There were previously 268 SARS-CoV-2 genomes available on GISAID where Pakistan was listed as the country of exposure with 90 assigned as B.1.1.7 or B.1.351 (Table 1; Supplementary Table S2). Most of these samples were associated with international travel from the country and were collected outside of Pakistan (Supplementary Table S2). A list of submitting authors of these data can be found in Supplementary Table S3. We combined all B.1.1.7 and B.1.351 public data with the genomes presented in this study to provide a snapshot of B.1.1.7 and B.1.351 dissemination (Supplementary Table S4).

**Table 1:** PANGO lineages of GISAID data where Pakistan is the country of exposure.

| Lineage | Count | Lineage | Count | Lineage | Count |
| --- | --- | --- | --- | --- | --- |
| B.1.1.7 | 87 | C.23 | 5 | B.1.525 | 1 |
| B.1.36 | 35 | B.1.36.24 | 3 | C.36 | 1 |
| B.1 | 30 | A | 3 | B.1.260 | 1 |
| B.1.471 | 24 | B | 3 | B.1.36.8 | 1 |
| B.1.1.1 | 18 | B.1.351 | 3 | B.1.523 | 1 |
| B.1.36.31 | 17 | C.11 | 2 | B.1.459 | 1 |
| B.6 | 12 | B.1.1.370 | 1 | B.1.562 | 1 |
| B.1.1 | 10 | B.1.36.22 | 1 | B.1.617.1 | 1 |
| B.1.36.34 | 5 | B.1.1.214 | 1 |  |  |

B.1.1.7 was likely introduced into Pakistan as the lineage emerged in the United Kingdom. B.1.1.7 samples are found in multiple clades spanning the entire phylogeny of B.1.1.7, suggesting the time to most recent common ancestor of all B.1.1.7 and B.1.1.7 in Pakistan was similar, which was calculated as October 2020 (Figure 1). The date of emergence of B.1.1.7 is before this date and has been calculated as September 2020 elsewhere (Galloway *et al*., 2021). These early introduction dates into Pakistan are plausible as the first confirmed B.1.1.7 genomes associated with Pakistan date back as early as late December 2020 (Figure 1).

**Figure 1:**
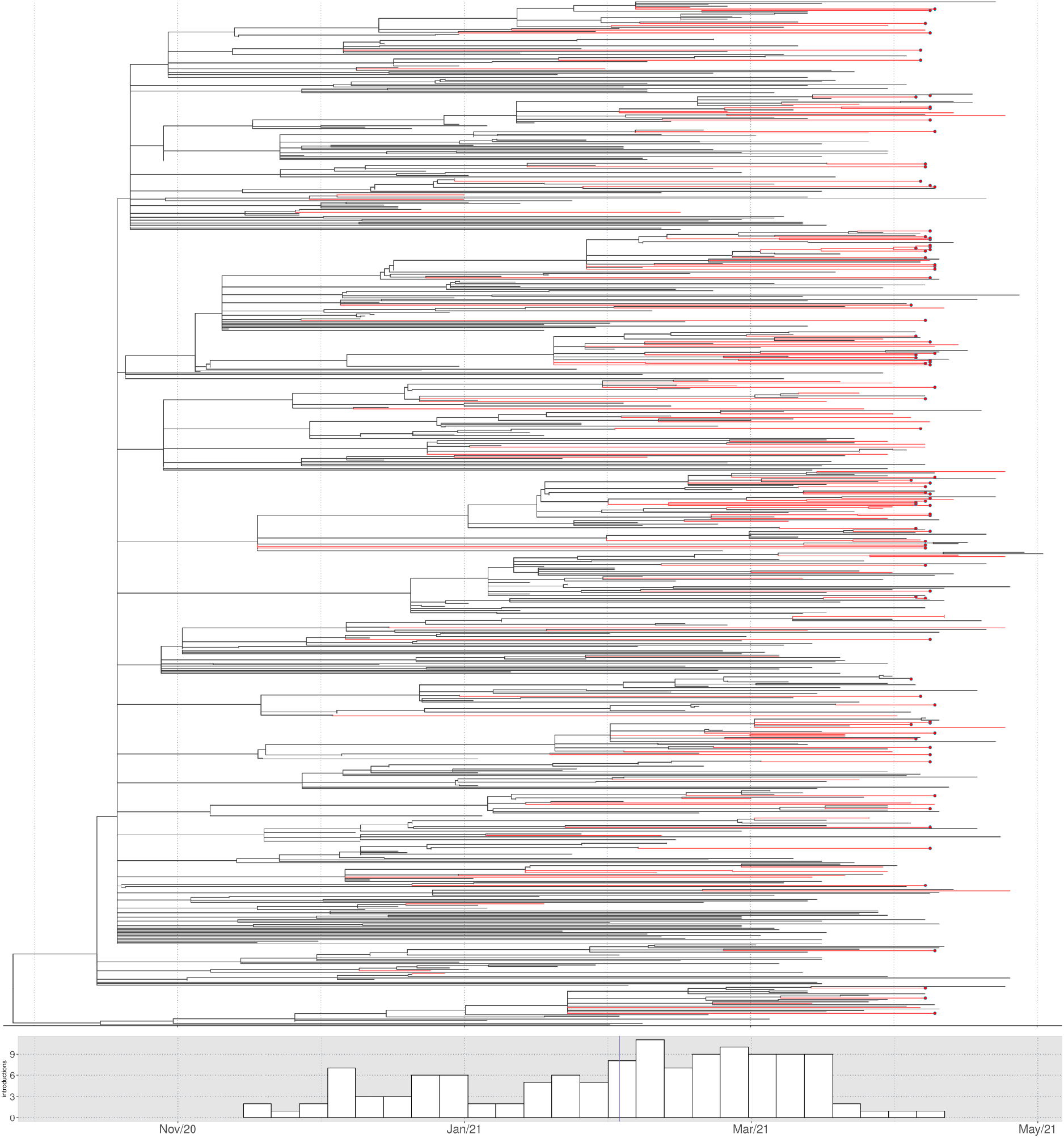
Phylogeny of B.1.1.7 including genomes from Pakistan. At the top, the maximum likelihood dated tree of lineage B.1.1.7, showing only samples close to sequences related to Pakistan. Branches are coloured red when all descendant tips have Pakistan as the country of exposure. Samples sequenced in the present study are highlighted as red dots. At the bottom we have the histogram of introductions into Pakistan over time, as estimated by ancestral state reconstruction, with the average time represented by a blue vertical line. Phylogenetic tree was estimated with IQTREE2 followed by divergence times estimation usingTreeTime after excluding outliers. The figure was plotted with ggtree and introduction events were estimated with castor.

By reconstructing the exposure history over the phylogenetic tree, we estimate the number of B.1.1.7 importations as 127 using the dated tree (Figure 1). More than half of the introductions into the country happened before March 2021, despite a constant rate of recent importations. Genomes from Pakistan were intermingled with genomes from elsewhere, as shown in Figure 2 where we calculated the patristic distance of genomes in this study to their closest neighbour and found that they were generally closer to samples from abroad. The number of substitutions to the nearest neighbour also varied (0-8 substitutions)(Figure 2). Considering that all samples in this study were sourced from patients with no travel history, this suggests that the current data does not fully capture transmission within Pakistan. The virus is detected and sequenced from the source (usually the United Kingdom) and then in some cases sampled in Pakistan immediately or after several months, where the number of substitutions were lower or higher respectively.

**Figure 2:**
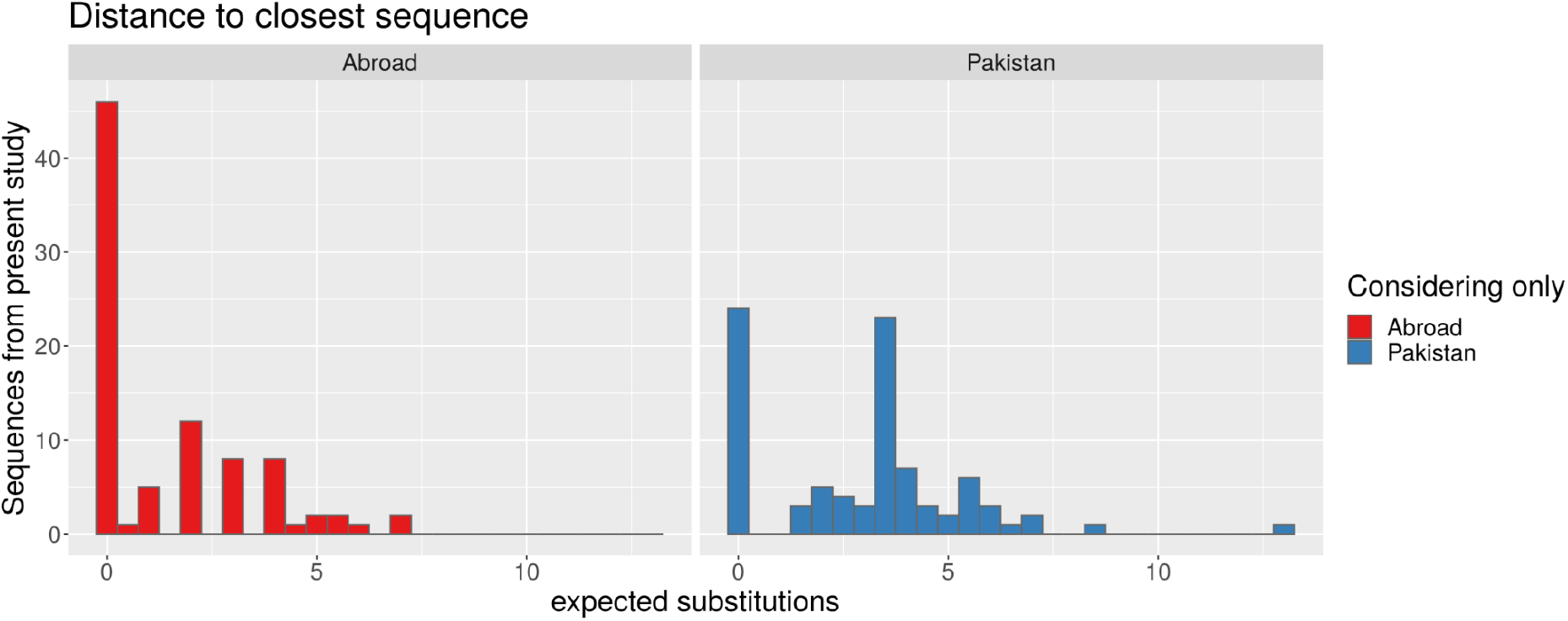
Distribution of distances to closest neighbours from Pakistan and abroad, for B.1.1.7 sequences. The number of expected substitutions is calculated from the patristic distances between leaves over the maximum likelihood tree, transformed from substitutions per site by multiplying by genome length. For each of the 88 UHS-PAK sequences, we find the closest distance considering only Pakistan (blue) or international (red) tips on the tree.

B.1.351 was likely introduced into Pakistan recently. B.1.351 samples were found in four clades in the phylogeny of B.1.351, suggesting at least 4 separate introductions into the country (Figure 3). The time to the most recent common ancestor of B.1.351 globally was calculated here as during September 2020, whereas the clades containing B.1.351 genomes from Pakistan date no earlier than February 2021 (Figure 3). The date of emergence of B.1.351 is before these dates and has been calculated as early August 2020 elsewhere (Tegally *et al*., 2021).

**Figure 3:**
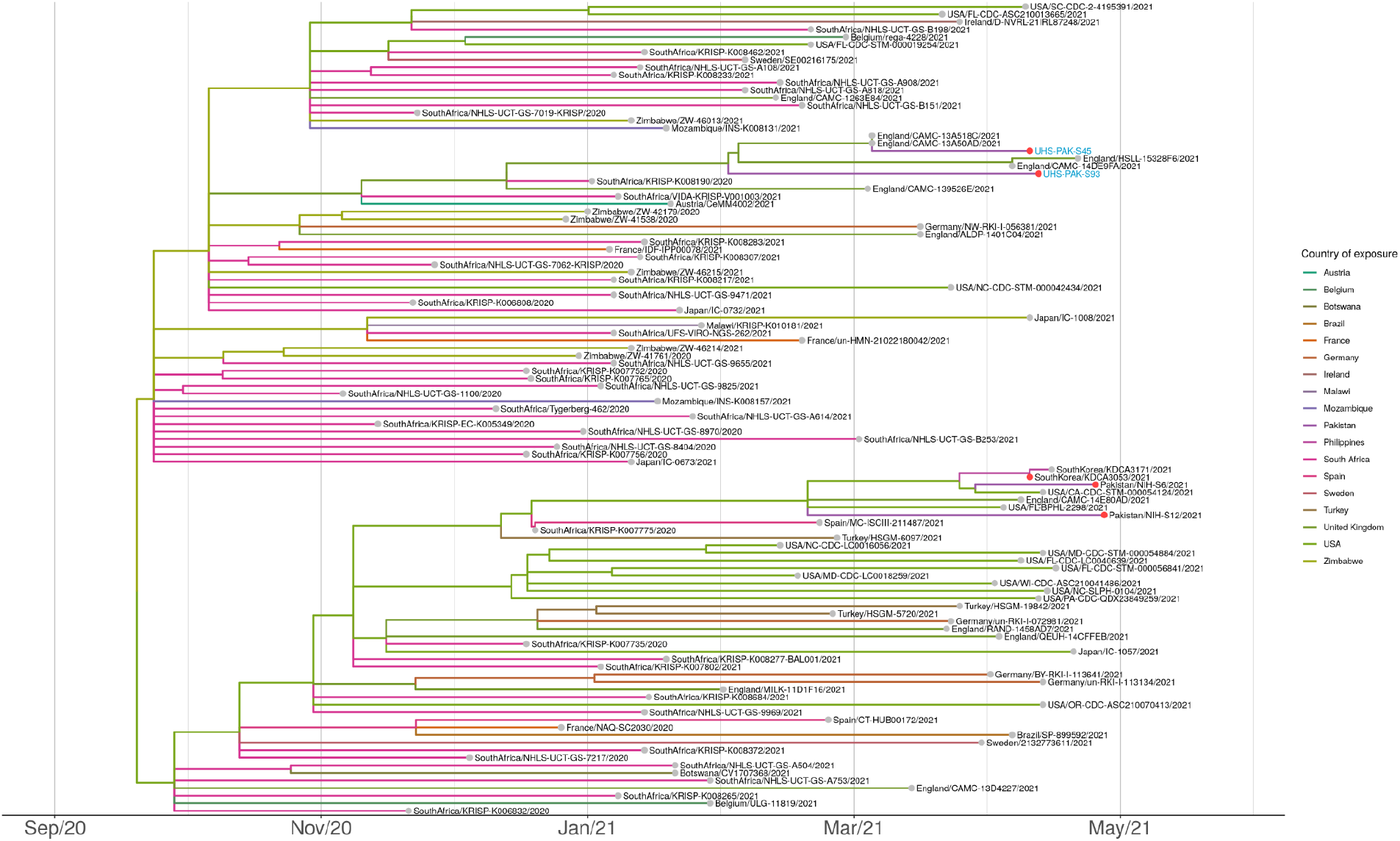
Maximum likelihood dated tree of lineage B.1.351 focused on samples sequenced in this study. Branches are coloured according to country of exposure, with Pakistan highlighted with a red circle. The two samples from the current study, UHS-PAK-S45 and UHS-PAK-S93, are labelled in blue.Phylogenetic tree was estimated with IQTREE2 (Minh *et al*., 2020) followed by divergence times estimation usingTreeTime after excluding outliers. The figure was plotted with ggtree (Yu *et al*., 2017).

It has been demonstrated (Abu-Raddad, Chemaitelly and Butt, 2021) that vaccines, in particular BNT162b2, are effective against B.1.1.7. Therefore, if this remains the dominant lineage in the region, public health vaccination policy can be implemented accordingly. However, whilst the prevalence of B.1.351, which has been linked to lower vaccine efficacy, is very low, this reinforces the need for prospective surveillance of SARS-CoV-2 using genome sequencing to inform public health interventions in a continual manner.

The most regular source of genomic sequencing is from travellers from Pakistan being sequenced by their destination countries, and being annotated as such in the public databases (GISAID). Japan is the largest contributor of genomes from travellers originating from Pakistan (111 out of 269). This indirect surveillance is useful but unreliable as travel restrictions and pre-flight testing can bias the results.

This report shows the critical importance of whole-genome sequencing of SARS-CoV-2 to determine the prevalence and changing epidemiology of different variants of the virus. This data is crucial to inform public health decision makers as well as to allow global epidemiology to be understood. Building capacity for sequencing and analysis of genomes in countries with high infection rates will be crucial for the global response to CoVID-19.

## Data Availability

Assembled/consensus genomes are available from GISAID (Shu and McCauley, 2017) subject to minimum quality control criteria. Raw reads are available from European Nucleotide Archive (ENA) in Bioproject PRJEB45462.

## Supplementary data

Supplementary table S1 - Samples sequenced in this study

Supplementary table S2 - GISAID metadata of samples associated with Pakistan

Supplementary Table S3 - GISAID Submitting author information

Supplementary Table S4 - GISAID metadata of B.1.351 and B.1.1.7

## Methods

### Genome sequencing and analysis

RNA was extracted using Viral RNA Extraction kit, FavorPrep™ and Polymerase Chain Reaction (RNA) was carried out using GenomeCoV19 Detection Kit by abm (cat: 628) on IQ 5 bioRad in diagnostic laboratories of Lahore Pakistan. Positive samples with a CT < 25 were randomly selected for genome sequencing. Viral RNA was converted in cDNA and was amplified using the ARTIC protocol v3 (LoCost) (Quick, 2020) with sequencing libraries prepared using CoronaHiT (Baker *et al*., 2021). We carried out genome sequencing using the Illumina NextSeq 500 platform.

The raw reads were demultiplexed using bcl2fastq (v2.20). The reads were used to generate a consensus sequence using the ARTIC bioinformatic pipeline (*Artic* Network, no date). Briefly, the reads had adapters trimmed with TrimGalore (Krueger, 2020) and were aligned to the WuhanHu-1 reference genome (accession MN908947.3) using BWA-MEM (v0.7.17) (Li, 2013); the ARTIC amplicons were trimmed and a consensus built using iVAR (v.1.2.3) (Grubaugh *et al*., 2019).

PANGO lineages assigned using Pangolin v2.4.2 and PangoLEARN model dated 2021-05-12 (Rambaut *et al*., 2020).

### Phylogenetic analysis

For the phylogenetic analysis, all sequences from GISAID where Pakistan is the country of exposure were downloaded and added to the sequences from the current study. All remaining sequences from GISAID were then compared to this data set where we kept the closest ones ―for each Pakistan sequence, the four closest from non-Pakistan were kept for subsequent analysis to provide context. The alignment and neighbour search were done with uvaia (de Oliveira Martins, Leonardo, 2021), and problematic (homoplasic or difficult to sequence) sites were masked from the alignment (Turakhia *et al*., 2020). For the B.1.1.7 and B.1.351 dated phylogenetic inference, we further enriched each alignment with more distant neighbours which maximised the phylogenetic diversity (Minh, Klaere and von Haeseler, 2006), based on the neighbour-joining tree (Simonsen and Pedersen, 2011) of all sequences close to each Pakistan sequence, for each lineage. Sequences with more than 10% N, or with incomplete date information were excluded from analysis. Clusters distant and unrelated to Pakistan sequences were reduced or removed by visual inspection of maximum likelihood trees.

From the 102 UHS-PAK sequences, 90 samples were included in the phylogenetic analysis: 88 from B.1.1.7 and 2 samples from B.1.351; the final alignments have 723 and 107 sequences, respectively. The maximum likelihood trees were inferred with IQTREE2 v2.1.2 (Minh *et al*., 2020) and the divergence times were estimated by marginalisation under a strict clock using TreeTime (Sagulenko, Puller and Neher, 2018). Ancestral state reconstruction of the country of exposure was done with castor (Louca and Doebeli, 2018) and trees were plotted with ggtree (Yu *et al*., 2017), both for R. The number of introductions into Pakistan was estimated by counting edges where the reconstructed probability of being exposed in Pakistan increased to one, weighting or not by the number of most parsimonious scenarios.

Due to unequal sampling and sequencing, the number of introductions is an underestimate and their dates are subject to selection bias (e.g. previous introductions were not sequenced due to regional differences or severity of infection).

## Ethics

This project was conducted under approval number UHS/REG-20/ERC/1758 from the University of Health Sciences Lahore Ethical Review Committee

